# A ReAct Agentic AI System for Natural Language Querying and Statistical Analysis of The Cancer Genome Atlas Clinical Data

**DOI:** 10.64898/2026.07.15.26358188

**Authors:** Rajashekar Korutla, Saeed Amal

## Abstract

The Cancer Genome Atlas (TCGA) holds clinical data for over 11,000 patients across 33 cancer types, but access is hard because of complex file structures, heterogeneous formats, and the need for programming. We present an agentic system for natural language querying and statistical analysis of TCGA clinical data. The system uses a large language model (LLM) as an autonomous Reasoning and Acting (ReAct) agent that selects from eight computational tools, including data extraction, descriptive statistics, Kaplan-Meier survival analysis with log-rank tests, hypothesis testing, and verification against the curated TCGA Pan-Cancer Clinical Data Resource (CDR). The agent reasons about intermediate results, adapts its approach, and returns clinically contextualized responses with source attribution and auditable traces. We introduce TCGA-Agent-Bench, 440 queries across five difficulty tiers with ground truth from the independently curated CDR, evaluated with dual metrics for numerical accuracy and clinical completeness. The system achieves 93.4% overall accuracy, outperforming a fixed rule-based pipeline (87.1%), a single-pass LLM (81.8%), and retrieval-augmented generation (66.9% on a 50-query subset). Most of the benchmark is answerable from the CDR alone, so we locate the extraction layer’s value in fields the CDR lacks, such as drug treatments, TNM components, and biomarkers: on 26 queries targeting these, the full system answers 100% versus 3.8% for a CDR-only configuration. A tool-based agentic architecture enables accurate, auditable natural language analysis of clinical repositories, with value driven by tool design and recovered fields rather than model scale.

## I. Introduction

The Cancer Genome Atlas (TCGA) is one of the largest cancer genomics and clinical data resources, covering over 11,000 patients across 33 cancer types [1], [2]. Its molecular data is widely used computationally, but the clinical data (demographics, diagnoses, treatments, survival) remains underused because clinical information is spread across many Extensible Markup Language (XML) and Portable Document Format (PDF) files per patient, encoded inconsistently, and requires programming to extract, link, and analyze [3].

Large language models (LLMs) understand and generate natural language well, creating new opportunities for data access [4], and agentic systems that autonomously plan and execute multi-step workflows while adapting to intermediate results have shown promise on complex analytical tasks [5], [6]. The Reasoning and Acting (ReAct) paradigm [7], where an LLM alternates reasoning traces with tool-calling actions, works well for multi-step decision-making, yet agentic systems with formal tool-use have not been applied to clinical data repositories like TCGA. Earlier TCGA-accessibility work centers on programmatic tools such as TCGA-Assembler [8] and TCGAbiolinks [9], which require coding, and web platforms like the UCSC Cancer Genomics Browser [11], which offer limited analytical flexibility. Recent LLM-based clinical-text systems [13], [14] focus on information extraction rather than interactive analytical querying. No existing system provides a natural language interface uniting autonomous data extraction, statistical analysis, and survival modeling in an agentic framework.

This paper makes four contributions. First, a data extraction pipeline covering all 33 TCGA cancer types that consolidates heterogeneous XML, JavaScript Object Notation (JSON), and PDF sources into structured records for 11,428 patients, parsing XML with namespace-aware traversal, handling institutional format variation, and resolving conflicts between initial and follow-up records by prioritizing the most recent data. This layer recovers many fields (drug treatments, tumor-node-metastasis (TNM) components, biomarker status, biospecimen metadata) absent from the curated TCGA Pan-Cancer Clinical Data Resource (CDR) [3], which let the agent answer questions a curated-reference-only system cannot. Second, an agentic architecture where an LLM autonomously selects from eight computational tools (including Kaplan-Meier (KM) survival analysis, log-rank tests, hypothesis testing, and a curated data verification tool) by reasoning about each query. Third, TCGA-Agent-Bench, a benchmark of 440 natural language queries across five difficulty tiers, with ground truth from the independently curated CDR [3]. Fourth, an evaluation using dual metrics (numerical accuracy and clinical completeness), comparison against three baselines (fixed pipeline, single-pass LLM, and retrieval-augmented generation (RAG) with embeddings), and ablation studies that justify each architectural component.

## II. Related Work

Extracting structured information from clinical data is well studied. Meystre et al. [15] reviewed clinical information extraction, noting rule-based approaches often outperform machine learning where domain knowledge can be encoded directly, and Jensen et al. [16] identified data heterogeneity and clinical language complexity as persistent barriers to electronic health record reuse. For TCGA, Liu et al. [3] developed the CDR, a standardized dataset of over 11,000 patients across 33 cancer types with curated endpoints for overall survival (OS), progression-free interval (PFI), disease-free interval (DFI), and disease-specific survival (DSS), now the accepted reference for validating extraction systems. Several tools ease retrieval: TCGA-Assembler [8], an R package automating downloading and processing; TCGAbiolinks [9] for integrative analysis and its Bioconductor workflows [10]; the UCSC Cancer Genomics Browser [11]; and cBioPortal [12], which offers visual exploration through predefined views. These improved accessibility, but the programmatic ones require coding and the visual portals expose fixed views rather than open-ended natural language analysis. Our system is complementary, targeting free-form analytical questions and returning computed statistics with reasoning traces.

Clinical natural language processing (NLP) has evolved rapidly with transformer-based models. Alsentzer et al. [17] showed domain-specific pre-training on clinical notes improves task performance; Kefeli and Tatonetti [18] used ClinicalBERT for cancer type classification on TCGA pathology reports, and Kefeli et al. [19] developed a model for TNM stage classification (area under the receiver operating characteristic curve 0.815 to 0.942). Singhal et al. [4] showed LLMs encode clinical knowledge but stressed the need for careful validation.

Agentic AI is an active area. Yao et al. [7] introduced ReAct, interleaving reasoning with actions to improve multi-step performance; it has been applied to code generation [20], scientific research [21], and database querying [22]. Tool-augmented LLMs are central: Schick et al. [23] showed incontext tool learning, and Patil et al. [24] demonstrated accurate application programming interface (API) call generation. Clinical tool-augmented approaches address electronic health record analysis and literature search [25], but not interactive querying of research repositories like TCGA. TCGA has also enabled extensive machine learning research: Linares-Blanco et al. [26] surveyed over 100 studies, finding Random Forest and Support Vector Machines prevalent and gene expression the most common modality; Mohammed et al. [27] reached 99.48% accuracy with ensemble deep learning on RNA-Seq; Thennavan et al. [28] extended histologic annotations for the TCGA breast cancer cohort; and Pan-Cancer analyses [1] and reviews [2] document TCGA’s structure across 12 and 30+ tumor types respectively.

## III. Methods

### A. System Architecture Overview

The system has three layers linked by an iterative reasoning loop (Fig. 1). A data extraction pipeline converts heterogeneous TCGA files (XML, JSON, PDF) into one structured JSON record per patient. Eight computational tools operate on those records: three retrieve data (single patient, single cohort, multiple cohorts), one queries the curated CDR reference, and four analyze (descriptive statistics, hypothesis testing, KM survival analysis, and data quality checking). A ReAct agent, an LLM, reads the natural language question, selects a tool, inspects the result, and decides whether to call another tool or write a final answer. The agent computes no statistics itself; all numerical work happens in the tools, and the model contributes language understanding, tool selection, and synthesis. Control passes between agent and tools until a response is produced, up to a fixed limit of 12 turns.

**Fig 1.**
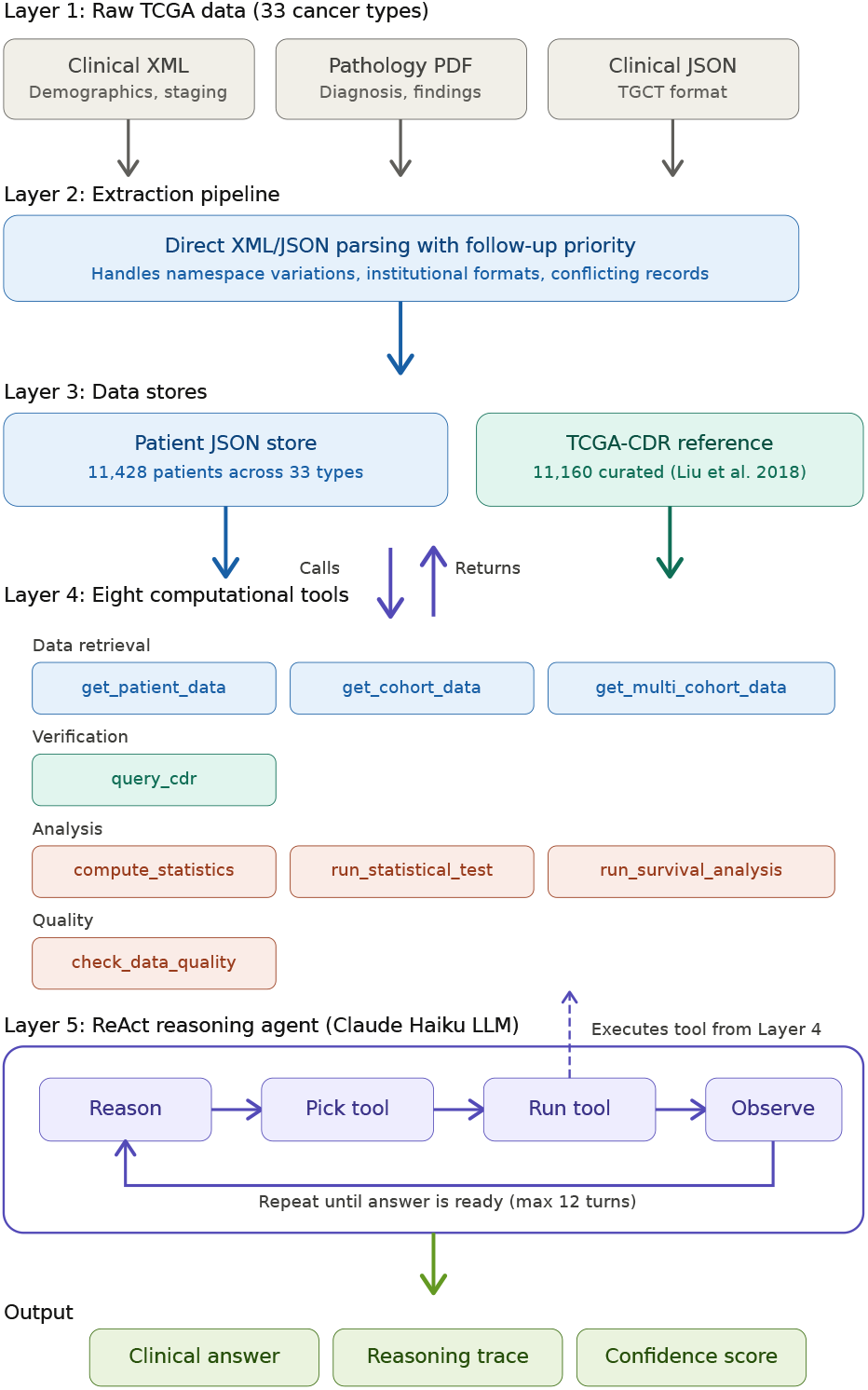
The system has three layers. The first is a data extraction pipeline that converts heterogeneous TCGA files into structured JSON records. The second consists of eight computational tools for data retrieval, curated data verification, statistical analysis, survival modeling, and data quality assessment. The third is a ReAct agent that uses an LLM to interpret queries, select and invoke tools, reason about intermediate results, and produce human-readable responses. These layers interact through an iterative reasoning loop.

### B. Data Extraction and Preprocessing

TCGA stores clinical information in multiple files per patient, often 20 or more: semi-structured XML (clinical, biospecimen, and supplemental site-specific factors from Nationwide Children’s Hospital or other institutions), unstructured PDF pathology reports, and genomic files. Two extraction challenges recur. First, vital status and survival data can appear in two locations within the clinical XML, the main patient section and separate follow-up sections: often the main section records an initial status (for example, “Alive”) while a later follow-up records an update (for example, “Dead” with days to death). In breast invasive carcinoma (BRCA) alone, 48 patients have conflicting vital status between sections; in stomach adenocarcinoma (STAD), 86; and in lung adenocarcinoma (LUAD), 61 of 188 deceased patients (32.4%) have death recorded only in follow-up, so an extractor reading only the main section would undercount deaths and inflate survival rates. Second, formats vary by cancer type: acute myeloid leukemia (LAML) stores clinical XML from a different institution (genome.wustl.edu rather than nationwidechildrens.org) for all 200 patients; testicular cancer (TGCT) provides clinical JSON for all 263 patients, 134 (51%) also having XML; and ovarian cancer (OV) is missing clinical XML for 21 of 608 patients (3.5%).

The pipeline parses XML with ElementTree using namespace-aware tag matching rather than flattening to text and applying regular expressions, matching elements by tag suffix (stripped of namespace prefixes) to handle namespace-Uniform Resource Identifier variation across institutions and TCGA versions. Clinical elements (demographics, staging, treatment, outcomes), biospecimen metadata (sample type, tumor nuclei and necrosis percentages), and supplemental annotations are extracted into a structured per-patient dictionary. The vital-status conflict is resolved by reading all followup sections, sorting by sequence number, and treating the most recent as authoritative: a final follow-up of “Dead” with a days to death value overrides an “Alive” main section. Institutional variation is handled with a broad file-matching pattern rather than institution-specific prefixes; for TGCT patients in JSON, a dedicated parser maps field names to the common schema. Patients with no clinical files are recorded as attempted-but-empty, letting the agent detect gaps and query the CDR as a fallback. PDFs are parsed with pdfplumber, preserving pathology-report tables in pipe-delimited format. Each patient’s JSON contains all clinical variables, drug treatments with start and end dates and therapy types, computed survival time (days converted to months, with censoring indicators), and extraction metadata. Across all 33 TCGA cancer types, the pipeline extracted data for 11,428 patients. The system does not impute missing values: each tool uses listwise deletion within the fields it needs and reports the number of patients used and per-field coverage, so reduced samples are visible rather than hidden.

We validated extraction against the CDR [3] across all overlapping fields. Vital status coverage exceeded 94% for 30 of 33 cancer types. Death counts matched CDR values closely: LUAD 188 versus 188, BRCA 150 versus 151, LAML 133 versus 133, and glioblastoma multiforme (GBM) 492 versus 489. This agreement holds at the individual-patient level, not only aggregate totals, so equal counts reflect the same patients rather than coincidence. Racial distributions agreed within 1 percentage point across all groups, and per-cancer-type patient counts differed by less than 5% from CDR values for 31 of 33 types.

The raw extraction and CDR play complementary roles. The CDR contains four curated fields the raw files do not expose in the same form: cause of death, a curated clinical stage, and the PFI event and time. The raw extraction, in turn, recovers fields the CDR does not contain at all: drug treatments with dates and therapy type (4,240 patients), pathologic TNM components (T for 7,888 patients, N for 7,840, M for 6,968), cancer-specific biomarkers such as estrogen receptor (ER), progesterone receptor (PR), and human epidermal growth factor receptor 2 (HER2) status in breast cancer and epidermal growth factor receptor (EGFR) and KRAS status in lung cancer, residual tumor status (4,447 patients), lymph nodes positive counts (3,950 patients), tobacco use history (2,927 patients), and biospecimen metadata such as tumor nuclei and necrosis percentages. These fields support treatment, staging, biomarker, and specimen questions that cannot be answered from the curated reference; a field-level summary is in Appendix F.

### C. Curated Data Reference (CDR Tool)

The system incorporates the CDR [3] as a queryable tool providing independently curated variables for 11,160 patients across all 33 cancer types: age, gender, race, pathologic stage, vital status, and four survival endpoints (OS, PFI, DFI, DSS). It supports single-patient lookup, cohort-level statistics, full-cohort retrieval, and verification of raw extraction against curated values (reporting matches, discrepancies, and fillable gaps). Raw extraction provides richer detail the CDR lacks; the CDR provides verified survival endpoints and fills gaps for patients with incomplete raw files. The agent decides at runtime which source to use and can cross-reference both.

### D. ReAct Agent Framework

The agent follows the ReAct paradigm [7] using the Anthropic Claude API [29] with native tool-use. Unlike fixed-sequence pipelines, it determines its strategy at runtime: each turn, the LLM receives the conversation history (prior reasoning, tool calls, results), generates a reasoning trace, and either invokes a tool or produces a final response, for up to 12 turns. A system prompt gives guidelines on tool selection, statistical reporting, and clinical interpretation without prescribing tool sequences; the agent reasons about the right approach per query. The full prompt and complete input and output schema for each of the eight tools are in Appendix A. The eight tools are summarized in Table I.

**TABLE I.** Summary of the eight computational tools available to the ReAct agent.

| Tool | Function |
| --- | --- |
| get_patient_data | Clinical data for a single patient |
| get_cohort_data | All patients in one cancer type |
| get_multi_cohort_data | Data across multiple cancer types |
| query_cdr | CDR lookup for verification and gap-filling |
| compute_statistics | Descriptive statistics, optional stratification |
| run_statistical_test | t-test, Mann-Whitney U, chi-square |
| run_survival_analysis | KM survival analysis with log-rank test |
| check_data_quality | Completeness and coverage gap reporting |

Tool choices follow the agent’s reasoning. Given “What is the survival rate for BRCA patients?”, it calls `get_cohort_data` (observing 1,098 patients), then `run_survival_analysis` and `compute_statistics`, and synthesizes simple rates and time-to-event estimates. For “Which of the 33 TCGA cancer types has the highest survival rate?”, it selects `get_multi_cohort_data` for all 33 types, then `compute_statistics` grouped by cancer type. Every step is logged for an auditable record. Hypothesis tests check assumptions rather than applying them blindly: before a two-group numerical comparison, `run_statistical_test` checks normality (Shapiro-Wilk) and equal variance (Levene), falling back from the t-test to Mann-Whitney U when violated and reporting which was used, and across multiple within-session comparisons it reports Bonferroni-adjusted p-values alongside raw values. Survival comparisons use KM with the log-rank test, which does not assume proportional hazards; for Cox extensions the tool can report a Schoenfeld residual diagnostic.

### E. Confidence Scoring

Each result includes a data-driven score summarizing the quality of the underlying data, not the model’s certainty. It combines three factors into a 0 to 100 score: sample size (0 to 40; 500+ patients scores 40, under 10 scores 5), coverage of the fields used (0 to 40), and vital-status coverage (0 to 20), categorized HIGH (85+), MODERATE (65 to 84), LOW (45 to 64), or VERY LOW (below 45). The weights and thresholds are heuristic and follow common clinical reporting conventions, not learned or calibrated, so we treat the score as a data-quality flag and return to its limitations in Sections IV-C and V-C.

### F. Benchmark Design (TCGA-Agent-Bench)

Ground truth comes from the CDR [3], an independently curated dataset with verified variables for 11,160 patients (age at diagnosis, gender, race, pathologic stage, vital status, overall survival time). Because ground truth is drawn from the CDR, the benchmark by construction tests only fields overlapping the CDR, not the additional extraction fields in Section III-B, a point we return to in the discussion. To ensure clean evaluation, the CDR tool was disabled during benchmarking; the agent answered using only raw extraction and its statistical tools. We designed 440 queries in five tiers of increasing difficulty: Tier 1 (50 single patient lookups), Tier 2 (115 single cohort statistics), Tier 3 (128 filtered subgroup analyses), Tier 4 (126 comparative analyses), and Tier 5 (21 multi-step cross-cancer ranking queries). Tier sizes follow the natural number of comparison units (a per-cancer-type statistic yields 33 queries) rather than an arbitrary target. Queries were generated programmatically from CDR fields using templated forms and checked by the authors for correctness, so they are more uniform in phrasing than free-form questions, a limitation examined in Section IV-G; full tiers and examples are in Appendix B. An author-constructed benchmark risks bias toward what the system does well, which three design aspects limit: ground truth is the independent CDR [3], not system output; tiers and sizes are fixed by data structure and cover every cancer type; and query forms target standard clinical quantities not specially tuned during development. These reduce but do not eliminate the bias; an independently authored query set is a natural next step.

We evaluate with two complementary metrics (full scoring rules in Appendices G and H). *Numerical accuracy* measures whether the agent produces the correct number, comparing extracted values against CDR ground truth within a tolerance of *±* 5% for rates (multiple extraction strategies), verifying each field for lookups, and scoring comparative queries with weighted partial credit for group identification, numerical proximity, and direction; rankings check whether the correct type appears. The tolerances reflect reporting granularity: 5 months for median survival and 5% for survival rates. Scoring is automated, and the authors spot-checked scored responses and found no discrepancies above tolerance. *Clinical completeness* measures methodological rigor on a 0 to 100 scale as a pattern-based proxy for good reporting, not human-judged quality. It is the fraction of applicable reporting components satisfied: six apply to every query (sample size reported; raw counts with percentages; proper survival methodology; dataquality or limitation acknowledgment; contextualized statistics; a cited source) and two more to comparative queries (direction of comparison; statistical significance), so non-comparative queries are scored out of six and comparative out of eight, with detection rules in Appendix G. Because these components describe capabilities the system was designed to exhibit, a high score reflects adherence to the intended reporting style rather than independent generalization. We report completeness across all 440 queries, and it is not validated against clinician ratings, which is left to future work.

### G. Baseline Designs

We compare three baselines, all with the same 11,428-patient dataset and statistical functions; only the reasoning mechanism varies. *Fixed Pipeline:* a rule-based parser detects cancer and query type from keywords, runs a hardcoded sequence (extract, compute, format) with no LLM, testing whether reasoning adds value over deterministic logic. *Single-Pass LLM:* the same functions compute all potentially relevant statistics, handed to the LLM in one prompt for one-shot interpretation, with no tool selection or loop. *RAG with Embeddings:* records are indexed in ChromaDB, retrieved per query by cosine similarity with cancer-type metadata filtering, statistics computed on retrieved records, and synthesized one-shot, testing whether autonomous tool selection beats retrieve-and-summarize.

### H. Implementation Details

The system is written in Python: NumPy and SciPy for statistics, lifelines for KM and log-rank tests, ChromaDB for the RAG baseline, and the Anthropic Claude API (`claude-haiku-4-5`) for reasoning, via Representational State Transfer with exponential backoff (up to 5 retries). All extraction and computation run locally; the LLM handles only interpretation, tool-selection reasoning, and synthesis, keeping results deterministic. Because the LLM samples responses, tool choices can vary slightly across runs, especially on multi-step queries (Section IV-E).

## IV. Results

### A. Data Extraction Coverage

The system processed all 33 TCGA cancer types, extracting 11,428 patients, with cohorts from 51 (cholangiocarcinoma, CHOL) to 1,098 (BRCA). Vital status coverage exceeded 94% for 30 of 33 types and per-type counts agreed with the CDR within 5% for 31 of 33. The slight excess over the CDR (11,428 versus 11,160) reflects patients present in raw files but excluded from the CDR on quality grounds, and every CDR patient is present in our extraction.

### B. Benchmark Performance

On all 440 TCGA-Agent-Bench queries (Table II), overall accuracy was 93.4%, with 361 queries (82.0%) perfect, 62 (14.1%) partial, and 17 (3.9%) zero. Repeating the full evaluation three times gave a stable 93.4% (standard deviation 0.2 points; 93.5%, 93.1%, 93.7%): variance was zero on Tiers 1 and 2, 0.6 points on Tier 3, 1.0 on Tier 4, and 2.2 on Tier 5, so variability is small and concentrated in multi-step queries where the tool-selection path can differ.

**TABLE II.** Benchmark performance by tier.

| Tier | Description | Queries | Accuracy | Perfect | Partial | Zero |
| --- | --- | --- | --- | --- | --- | --- |
| 1 | Single patient lookup | 50 | 100.0% | 50 | 0 | 0 |
| 2 | Cohort statistics | 115 | 99.1% | 114 | 0 | 1 |
| 3 | Filtered subgroup | 128 | 87.5% | 109 | 6 | 13 |
| 4 | Comparative analysis | 126 | 92.8% | 70 | 56 | 0 |
| 5 | Cross-cancer ranking | 21 | 85.7% | 18 | 0 | 3 |
| **Total** |  | **440** | **93.4%** | **361** | **62** | **17** |

Accuracy fell as tiers grew harder (Table II). Tier 1 reached 100% (age, gender, and vital status correct for all 50 patients, each in 2 turns); Tier 2 reached 99.1% (114 of 115, including all 33 survival-rate and all 33 average-age queries, one distribution query failing on formatting); Tier 3 reached 87.5%, with its 13 zero-score queries concentrated in median survival (9) and filtered subgroup survival (4) where raw-versus-CDR coverage differences exceeded tolerance; Tier 4 reached 92.8%, all 126 earning at least partial credit (70 perfect, 56 partial at typically 0.6 to 0.9) with correct groups and directions; and Tier 5 reached 85.7% (18 of 21), correctly identifying the highest survival rate (prostate adenocarcinoma, PRAD), lowest (mesothelioma, MESO), youngest mean age (TGCT), and highest male proportion (TGCT), with three reaching the turn limit. Of the 17 zero-score queries overall, 9 involve KM median survival diverging from the CDR beyond the 5-month tolerance, 4 filtered survival rates with coverage differences beyond 5%, 3 Tier 5 queries reaching the 12-turn limit, and 1 a formatting mismatch (Appendix E). No query produced a wrong analytical conclusion, and in all 126 comparative queries the agent identified the correct direction even when exact numbers received only partial credit.

### C. Confidence Scoring

Confidence scores were computed for all 440 queries (mean 93.8 of 100, median 95.0): 416 (94.5%) high and 24 (5.5%) moderate, none low or very low. Because the score was earlier framed as calibrated, we tested that directly: standard summaries are consistent with a reasonable data-quality signal (expected calibration error 0.06, Brier score 0.047), but the rank correlation with per-query accuracy is effectively zero (Spearman *ρ* = *−* 0.02, not significant) and selective-prediction accuracy is flat (93.1% high versus 92.5% moderate), as expected when almost all queries fall in the high band. We therefore do not claim the score predicts correctness; it is a data-quality flag, and learned outcome-aware calibration is future work.

### D. Baseline Comparison

We compared the ReAct agent against the fixed pipeline and single-pass LLM on the full 440 queries, and against RAG on a representative 50-query subset (10 per tier). RAG is reported on the subset because full-cohort re-indexing was unreliable in our environment, and its subset accuracy is low enough not to change the ranking. All use the same data and statistical functions (Table III).

**TABLE 3.** Baseline comparison. React, Fixed Pipeline, and Single-Pass LLM are evaluated on the full 440-query benchmark; RAG is evaluated on a 50-query subset (10 per tier).

| Approach | Accuracy | Evaluation set |
| --- | --- | --- |
| RAG + Stats | 66.9% | 50-query subset |
| Single-Pass LLM | 81.8% | full 440 |
| Fixed Pipeline | 87.1% | full 440 |
| ReAct Agent | 93.4% | full 440 |

The ReAct agent (93.4%) outperforms the fixed pipeline (87.1%) and single-pass LLM (81.8%), with differences small on simple tiers and largest on comparative and ranking tiers; on Tier 4 the agent leads the nearest baseline by about 30 points. This also separates the extraction layer from the reasoning loop: the Fixed Pipeline uses the same extraction and functions without LLM reasoning, so its 87.1% reflects extraction and tools alone, and the 6.3-point gap to 93.4% (about 30 points on Tier 4) isolates the reasoning loop’s contribution. RAG performs worst (66.9%): semantic retrieval optimizes for relevance, not completeness, so it may return a non-representative subset and produce incorrect statistics, a structural weakness for population-level queries not resolved by a better embedding model.

### E. How Much of the Benchmark the Curated Reference Alone Can Answer

Because ground truth comes from the CDR, we asked how much a CDR-only configuration (using the reference for retrieval instead of raw extraction) could answer. On the full 440 queries it reached 91.6%, close to the full system’s 93.4%. Neither reaches 100% despite CDR-derived ground truth because answers are computed by tools (for example, a KM median) and scored by pattern matching within tolerance rather than looked up, so small computation or phrasing differences cost both points. This means the curated reference alone answers most queries, so the benchmark does not by itself show the extraction layer improves accuracy, because it can only ask about fields the CDR already contains. The extraction layer’s distinct value is in the fields the CDR does not contain (Section III-B and Appendix F), demonstrated directly next. Multi-step Tiers 4 and 5 show run-to-run variation from sampled tool selection while single-record and single-cohort tiers are stable.

### F. Direct Test of the Extraction Layer on Non-Reference Fields

To measure the extraction layer’s contribution directly, we built 26 queries about fields the CDR does not contain: biomarker status (ER, PR, HER2 in breast cancer), pathologic TNM staging, administered drugs, and residual tumor status. Ground truth is computed from the extracted records, relying on the extraction having been validated on overlapping fields (Section III-B). We ran both the full agent and the CDR-only configuration (Table IV).

**TABLE IV.** Accuracy on 26 queries targeting fields absent from the curated reference. The CDR-only configuration cannot answer these queries because the fields do not exist in the CDR.

| Field category | Queries | Full agent | CDR-only |
| --- | --- | --- | --- |
| Biomarker (ER/PR/HER2) | 4 | 100.0% | 0.0% |
| TNM staging | 10 | 100.0% | 10.0% |
| Drugs | 10 | 100.0% | 0.0% |
| Residual tumor | 2 | 100.0% | 0.0% |
| **Total** | **26** | **100.0%** | **3.8%** |

The full agent answered all 26 correctly, while CDR-only reached 3.8% (one TNM query, from a related curated stage field). The 96-point gap is not about reasoning: the required fields do not exist in the reference, so a CDR-limited system cannot answer them at all. This is the concrete demonstration behind Section III-B: the extraction layer makes treatment, staging, biomarker, and specimen questions answerable in the first place. Ground truth here derives from the same extraction the agent queries, so this measures correct retrieval and computation over these fields, not extraction correctness itself, which is established in Section III-B.

### G. Robustness to Query Phrasing

Because the benchmark queries are templated, we tested wording dependence. We selected eight query types spanning Tiers 2 through 5 and wrote two to three natural paraphrases of each with identical ground truth (for example, “What is the survival rate for LUAD patients?” reworded as “For lung adenocarcinoma patients, what fraction are still alive?”), yielding 17 paraphrases. On the eight matched templated queries the agent scored 100%; on the 17 paraphrases, 88.9%. Accuracy held at 100% for cohort-statistic and cross-cancer ranking paraphrases, while drops concentrated in one comparative query (partial credit, direction still correct) and one filtered-subgroup query where indirect phrasing (“late-stage (Stage IV)”) moved the result outside tolerance. The system is largely robust to natural phrasing, with residual sensitivity on indirectly-filtered subgroups. These author-written paraphrases are modest in number, so this is indicative rather than a large-scale study.

### H. Ablation Study

Removing one tool at a time on the 50-query subset with both metrics (Table V), the reasoning loop is most impactful, its removal cutting accuracy 9.1 points and completeness 22.0 points, concentrated in Tier 4 (89.3% to 53.6%). The multi-cohort tool causes the second-largest completeness drop (12.4 points), with Tier 5 falling from 80% to 60%. Two tools (KM survival, statistical testing) show small accuracy gains when removed (+3.4% and +2.2%) but substantial completeness losses (*−* 10.8 and*−* 9.9 points), because removing them yields simpler responses that are easier to score numerically while the richer responses they enable (time-to-event analysis, dataquality caveats, source cross-referencing) are not captured by accuracy alone. Removing the data-quality tool has minimal accuracy impact (*−* 0.6%) but reduces completeness 8.0 points as the agent stops flagging coverage gaps and small samples. Across 440 queries the agent averaged 3.2 tool calls per query, consistently choosing `run_survival_analysis` for time-to-event questions and `compute_statistics` for distributions, and calling `check_data_quality` unprompted on low-coverage cohorts (Appendix C). The full run took 4,523 seconds (about 75 minutes), averaging 10.3 seconds per query (2 to 4 seconds for lookups, 5 to 15 for single-cohort analyses, 30 to 120 for pan-cancer ranking) at roughly USD 5.35 total, about USD 0.012 per query; the bottleneck for complex queries is data loading, not LLM inference. As a scale check, a pan-cancer racial-distribution analysis across all 33 types completed in 3 turns and 36.6 seconds, agreeing with CDR values within 1 percentage point for every group (Appendix D).

**TABLE V.** Ablation results with dual metrics of accuracy and completeness on the 50-query subset.

| Approach | Acc. | Compl. | Acc $\Delta$ | CC $\Delta$ |
| --- | --- | --- | --- | --- |
| Full Agent (7 tools) | 89.9% | 44.8 | — | — |
| Remove KM survival | 93.2% | 33.9 | +3.4% | −10.8 |
| Remove multi-cohort | 88.9% | 32.3 | −1.0% | −12.4 |
| Remove data quality | 89.2% | 36.8 | −0.6% | −8.0 |
| Remove stat tests | 92.1% | 34.8 | +2.2% | −9.9 |
| No reasoning loop | 80.7% | 22.8 | −9.1% | −22.0 |

## V. Discussion

### A. Advances Over Prior Approaches

Our system differs from earlier TCGA access tools beyond architecture: pipeline-based systems follow fixed sequences, while our ReAct agent decides its strategy per query. Three behaviors support the agentic claim: it selects different tool combinations and sequences by query, chosen by the LLM at runtime rather than hardcoded; it adjusts to intermediate results, calling the quality tool on low coverage and the CDR as a fallback when extraction is incomplete; and its reasoning traces make the process transparent, showing not just the answer but how it was reached.

### B. Evaluation Findings

Three findings stand out. First, a single accuracy metric is insufficient: removing the KM tool raises accuracy 3.4% while cutting completeness 10.8 points, so an accuracy-only evaluation would prefer the thinner response. The dual-metric approach exposes this, with completeness measuring whether a response provides what a clinical researcher needs (sample sizes, proper methodology, limitations, source attribution); it is a pattern-based proxy, and validating it against clinician ratings is an important next step. Second, RAG’s low score (66.9%) is specific to statistical queries: semantic retrieval optimizes for relevance, not completeness, so asked for the survival rate of bladder urothelial carcinoma (BLCA) patients it may omit some of the 412 patients, and statistics on an incomplete sample are wrong regardless of retrieval quality. Where the answer depends on complete aggregation rather than finding the right document, retrieval is structurally disadvantaged. Third, the reasoning loop earns its cost on complex queries: all approaches perform equally on simple lookups, but comparative analysis and cross-cancer ranking require planning and adapting midway, which one-shot approaches cannot do.

An honest reading separates two questions. Can an agent answer natural language statistical questions over TCGA using only extracted data? Yes, at 93.4%. Does raw extraction improve accuracy over the CDR on this benchmark? Largely not, because a CDR-only configuration reaches 91.6% and the benchmark only asks about CDR fields. We therefore locate the extraction layer’s value in the treatment, staging, biomarker, and specimen fields the CDR lacks (Appendix F): our focused experiment (Section IV-F) shows the full system answering all 26 non-reference queries while CDR-only answers almost none, a 96-point gap arising purely from field absence. A benchmark drawn from a curated reference therefore understates the value of richer extraction, and a separate evaluation on non-reference fields is needed to reveal it. Because the CDR is both a system tool and the ground-truth source, we disabled it during evaluation, so the 93.4% reflects independent extraction and analysis; the CDR tool remains in the full system for real-world use, where cross-verification adds clinical value. Finally, although general-purpose agentic coding assistants can now assemble analysis pipelines from natural language, we see this work as complementary: the non-trivial parts are the TCGA-specific extraction pipeline (institutional format variation, follow-up conflicts, mixed XML, JSON, and PDF across 33 types), the verifiable 440-query benchmark with independent ground truth, and the dual-metric framework, all of which a general assistant would still need to produce trustworthy, checkable results.

### C. Limitations and Future Directions

Several limitations point to future work. The benchmark is templated, so its queries are more uniform than free-form questions; clinician-authored queries are a planned extension. It draws ground truth from the CDR and so does not measure the accuracy contribution of the extraction layer’s additional fields; a benchmark targeting treatment, biomarker, and staging fields would close this gap. Multi-step queries show run-to-run variation from sampled tool selection; reporting means and variability across repeated runs, which we recommend for agentic evaluations generally, gives a fuller picture. The confidence score is a heuristic data-quality flag that does not predict correctness; learned, outcome-aware calibration with reliability curves and selective prediction is future work. Clinical completeness reflects an intended reporting style rather than expert-judged correctness, so clinician validation is needed. The evaluation here is quantitative; studying real use by clinician-researchers is a natural direction. XML-parsing extraction restricts the system to explicitly recorded information, so adding clinical NLP for entity recognition from unstructured pathology reports would broaden coverage, and the agent does not yet self-verify against expected ranges or process data in parallel, so parallelized extraction would speed pan-cancer queries.

We deliberately used a small, low-cost model (`claude-haiku-4-5`). Because the architecture delegates every numerical operation to deterministic tools and asks the model only to interpret, select tools, and synthesize, a small model suffices and 93.4% is achieved without a frontier model, supporting our claim that value comes from tool design rather than model scale; a larger model is a drop-in replacement that might further reduce variation on the hardest tiers. Because the reasoning component is a hosted model updated over time, its behavior can drift when the version changes, but deterministic local tools produce all numerical results, so drift can affect tool selection and phrasing but not computed statistics, and the reasoning traces make any change visible. Pinning a model version and periodically re-running the benchmark would let a deployment monitor drift, which we recommend for production.

Finally, the system currently handles only clinical data. TCGA directories also contain RNA-seq expression, somatic mutation calls, copy number variation, DNA methylation, microRNA expression, protein levels, and whole-slide histology images. The tool-based architecture is extensible: adding a modality requires new tools, not redesigning the agent, enabling integrated genomic-clinical queries such as “What mutations are associated with poor survival in Stage III LUAD?” without changing the framework. Other extensions include Genomic Data Commons (GDC) API integration, Cox proportional-hazards and competing-risks survival methods, visualization, and web deployment.

## VI. Conclusion

We present a ReAct agentic system for natural language querying and statistical analysis of TCGA clinical data across all 33 cancer types and 11,428 patients. With the CDR tool disabled during evaluation, it achieves 93.4% accuracy on TCGA-Agent-Bench, a 440-query benchmark validated against the CDR, outperforming a fixed pipeline (87.1%), a single-pass LLM (81.8%), and a RAG baseline (66.9% on a subset), with the largest gains on complex comparative and ranking queries. We show transparently that a CDR-only configuration also answers most of the benchmark (91.6%), and we locate the extraction layer’s value in the treatment, staging, biomarker, and specimen fields the curated reference lacks; on 26 queries targeting those fields, the full system answers 100% while CDR-only answers 3.8%. Repeating the full benchmark three times gave a stable 93.4% (standard deviation 0.2 points), with variability confined to the hardest multi-step tiers. Ablation studies using dual metrics justify every tool, showing that tools which marginally affect accuracy still contribute substantially to clinical rigor.

The main contributions are the extraction pipeline that handles institutional format variations and follow-up conflicts across 33 cancer types and recovers many fields absent from the curated reference; the eight-tool agentic architecture with transparent reasoning traces; the TCGA-Agent-Bench benchmark with 440 queries across five tiers; and the dual-metric evaluation framework. As clinical data repositories grow in size and complexity, agentic systems like this one can turn stored data into research insights accessible to both computational and clinical researchers. The architecture applies beyond TCGA: any repository with distributed, heterogeneous storage could benefit from an agentic querying approach, and by recording every reasoning step, tool choice, and intermediate result, the system lets researchers audit the analytical process, a necessary property for responsible use of AI in clinical settings.

## Data Availability

All data underlying this study are derived from The Cancer Genome Atlas (TCGA), which is publicly available through the Genomic Data Commons (GDC) Data Portal (https://portal.gdc.cancer.gov/). The TCGA Pan-Cancer Clinical Data Resource (CDR) used as ground truth is available as supplemental material to Liu et al. (2018), Cell 173(2):400-416 (https://doi.org/10.1016/j.cell.2018.02.052). The TCGA-Agent-Bench benchmark (the 440 queries with their CDR-derived ground truth), the non-reference-field benchmark, and the evaluation and scoring code will be released in a public repository (GitHub) upon publication. The structured patient records analyzed by the system are derived from the public TCGA sources above and can be regenerated using the described extraction pipeline they are not redistributed separately.

https://portal.gdc.cancer.gov/

## Appendix A Tool Definitions

The agent can call eight computational tools. For each tool we give its inputs, its outputs, and the reasoning behind its design.

- `get_patient_data`: Extracts clinical data for a single patient by barcode identifier from the structured JSON store. Returns demographics, diagnosis, staging, biomarkers, drug treatments, and outcome data.
- `get_cohort_data`: Extracts clinical data for all patients in a single cancer type, storing results in session memory for subsequent analysis. Returns an extraction summary with patient count and available fields.
- `get_multi_cohort_data`: Extracts data across multiple cancer types at once, enabling pan-cancer analyses. Aggregates patients from up to all 33 types in a single call, which is needed for cross-cancer comparisons and ranking queries.
- `query_cdr`: Queries the CDR curated reference. Supports patient lookup, cohort statistics, full cohort retrieval, and verification of raw extraction against curated values. The agent uses this to fill gaps when raw files are incomplete or to cross-check critical fields like vital status.
- `compute_statistics`: Calculates descriptive statistics on the loaded cohort: mean, median, and standard deviation for numerical fields; frequency distributions for categorical fields; and alive and dead survival rates. A `groupby` parameter enables stratified analysis, for example by race, gender, stage, or cancer type.
- `run_statistical_test`: Runs hypothesis tests including the independent samples t-test, Mann-Whitney U test, and chi-square test. Before a two-group numerical comparison, it checks normality (Shapiro-Wilk) and equal variance (Levene) and falls back from the t-test to Mann-Whitney U when assumptions are violated, reporting which test was used. When several comparisons are run within one session, it reports Bonferroni-adjusted p-values alongside raw values. Returns test statistics, p-values, adjusted p-values where applicable, and significance indicators.
- `run_survival_analysis`: Performs KM survival analysis using the lifelines library. Reports median survival time, survival probabilities at 12, 36, and 60 months, and event and censoring counts. Supports stratification by any categorical variable, with automatic log-rank test computation for two-group comparisons. The log-rank test does not assume proportional hazards; for Cox model extensions the tool can report a Schoenfeld residual diagnostic.
- `check_data_quality`: Reports data completeness for specified fields, including present and missing counts, coverage percentages, and warnings for fields below 50% coverage or cohorts under 30 patients.

## Appendix B Benchmark Query Construction

We designed 440 natural language queries in five tiers of increasing difficulty. Tier sizes follow the natural number of comparison units (for example, one query per cancer type yields 33 queries) rather than a fixed target. Queries were generated from templated question forms populated with CDR fields, then checked by the authors for ground-truth correctness. The queries are therefore more uniform in phrasing than free-form user questions; collecting paraphrased and clinician-authored queries is planned future work.

- **Tier 1, Single Patient Lookups (50 queries):** Direct retrieval of demographic and clinical fields for individual patients by TCGA barcode. These test basic extraction accuracy. Example: “What is the vital status and age at diagnosis of patient TCGA-A2-A0T1?”
- **Tier 2, Single Cohort Statistics (115 queries):** Population-level analyses within one cancer type, including survival rates for all 33 types, average age for all 33 types, gender distributions for 20 types, stage distributions for 15 types, and racial distributions for 15 types. Example: “What is the overall survival rate for LUAD patients?”
- **Tier 3, Filtered Subgroup Analysis (128 queries):** Analyses that require filtering before computation: survival by stage, average age by gender, survival by race, median survival for deceased patients, and survival by age group. Example: “What is the median survival for deceased Stage IV STAD patients?”
- **Tier 4, Comparative Analysis (126 queries):** Multi-group comparisons including cross-cancer survival, alive versus deceased characteristics, survival by race within cancer types, survival by gender, Stage I versus Stage IV comparisons, and young versus old patient comparisons. Example: “Compare survival rates between BRCA and PCPG patients.”
- **Tier 5, Multi-step Ranking (21 queries):** Queries requiring analysis across multiple or all 33 cancer types to identify extremes, rank cancer types by metrics, or compute pan-cancer distributions. Example: “Which of the 33 TCGA cancer types has the highest survival rate?”

## Appendix C Agent Behavior Analysis

### Reasoning Patterns

Analysis of reasoning traces across 440 evaluation queries (CDR tool disabled) shows consistent and appropriate strategies. For survival queries, the agent follows a three-turn sequence: extract cohort data, run survival analysis, then synthesize a response with both simple rates and KM estimates. Comparative queries typically use four turns, adding grouped statistics or statistical testing. Ranking queries across all 33 types use `get_multi_cohort_data` to load all data in one call, then apply `compute_statistics` with the appropriate grouping. In production use with the CDR tool enabled, the agent may add verification turns to cross-reference extracted values against curated data.

### Tool Utilization

Across 440 queries, the agent averaged 3.2 tool calls per query. The most frequently used tools were `get_cohort_data`, `compute_statistics`, `run_survival_analysis`, and `get_multi_cohort_data`. The agent consistently chose `run_survival_analysis` for time-to-event questions and `compute_statistics` for distribution and count-based questions, showing context-appropriate tool selection.

### Adaptive Behavior

The agent showed adaptive behavior in several cases. When it encountered cohorts with low data coverage, it called `check_data_quality` on its own before reporting results. When computing survival for filtered subgroups, for example Stage III within a cancer type, it loaded the full cohort first, then applied filtering within its statistical analysis rather than filtering during extraction.

## Appendix D Pan-Cancer Racial Distribution Analysis

The agent loaded 11,428 patients through `get_multi_cohort_data`, computed racial distributions with `compute_statistics`, and checked data quality, completing the analysis in 3 turns and 36.6 seconds. Table VI shows the overall racial distribution compared against CDR reference values. Agreement was strong across all categories, with absolute differences below 1 percentage point for every group, validating the extraction pipeline for demographic fields.

**TABLE VI.** Overall racial distribution: system extraction versus CDR ground truth.

| Race | System<br>(n=11,428) | CDR<br>(n=11,160) |
| --- | --- | --- |
| White | 8,456 (83.1%) | 8,186 (82.9%) |
| Black or African American | 985 (9.7%) | 934 (9.5%) |
| Asian | 685 (6.7%) | 675 (6.8%) |
| American Indian or Alaska Native | 28 (0.3%) | 27 (0.3%) |
| Native Hawaiian or Other Pacific Islander | 13 (0.1%) | 13 (0.1%) |
| Missing or Not Available | 1,261 (11.0%) | 1,193 (10.7%) |

Table VII shows the racial composition by cancer type. White%, Black%, and Asian% are computed among patients with available race data. Coverage% indicates the proportion with non-missing race information. Black or African American patients have their highest representation in kidney renal papillary cell carcinoma (KIRP, 22.1%), colon adenocarcinoma (COAD, 20.7%), and uterine corpus endometrial carcinoma (UCEC, 21.1%), but are nearly absent from uveal melanoma (UVM, 0.0%) and skin cutaneous melanoma (SKCM, 0.2%). Asian patients are most represented in liver hepatocellular carcinoma (LIHC, 43.9%), lymphoid neoplasm diffuse large B-cell lymphoma (DLBC, 37.5%), and esophageal carcinoma (ESCA, 27.9%), consistent with the higher incidence of these cancers in Asian populations. Data completeness varies widely, from 100% in MESO to 49.0% in TGCT. The overall TCGA cohort is predominantly White (83.1%), substantially overrepresenting this group relative to the U.S. population (approximately 60% non-Hispanic White per Census data), a well-known limitation with implications for health disparities and population-specific biomarker research.

**TABLE VII.** Racial distribution by cancer type.

| Cancer | N | White | White% | Black | Black% | Asian | Asian% | Missing | Cov.% |
| --- | --- | --- | --- | --- | --- | --- | --- | --- | --- |
| ACC | 92 | 78 | 96.3 | 1 | 1.2 | 2 | 2.5 | 11 | 88.0 |
| BLCA | 412 | 327 | 83.0 | 23 | 5.8 | 44 | 11.2 | 18 | 95.6 |
| BRCA | 1098 | 757 | 75.5 | 183 | 18.3 | 61 | 6.1 | 96 | 91.3 |
| CESC | 307 | 211 | 77.9 | 30 | 11.1 | 20 | 7.4 | 36 | 88.3 |
| CHOL | 51 | 41 | 87.2 | 3 | 6.4 | 3 | 6.4 | 4 | 92.2 |
| COAD | 461 | 214 | 75.1 | 59 | 20.7 | 11 | 3.9 | 176 | 61.8 |
| DLBC | 58 | 29 | 60.4 | 1 | 2.1 | 18 | 37.5 | 10 | 82.8 |
| ESCA | 185 | 114 | 69.1 | 5 | 3.0 | 46 | 27.9 | 20 | 89.2 |
| GBM | 617 | 507 | 88.8 | 51 | 8.9 | 13 | 2.3 | 46 | 92.5 |
| HNSC | 528 | 452 | 88.1 | 48 | 9.4 | 11 | 2.1 | 15 | 97.2 |
| KICH | 113 | 95 | 85.6 | 12 | 10.8 | 4 | 3.6 | 2 | 98.2 |
| KIRC | 537 | 466 | 87.9 | 56 | 10.6 | 8 | 1.5 | 7 | 98.7 |
| KIRP | 291 | 207 | 75.0 | 61 | 22.1 | 6 | 2.2 | 15 | 94.8 |
| LAML | 200 | 181 | 91.4 | 15 | 7.6 | 2 | 1.0 | 2 | 99.0 |
| LGG | 516 | 475 | 94.1 | 21 | 4.2 | 8 | 1.6 | 11 | 97.9 |
| LIHC | 377 | 187 | 51.0 | 17 | 4.6 | 161 | 43.9 | 10 | 97.3 |
| LUAD | 585 | 393 | 86.4 | 53 | 11.6 | 8 | 1.8 | 130 | 77.8 |
| LUSC | 504 | 351 | 89.8 | 31 | 7.9 | 9 | 2.3 | 113 | 77.6 |
| MESO | 87 | 85 | 97.7 | 1 | 1.1 | 1 | 1.1 | 0 | 100.0 |
| OV | 608 | 498 | 89.6 | 34 | 6.1 | 20 | 3.6 | 52 | 91.4 |
| PAAD | 185 | 162 | 90.0 | 7 | 3.9 | 11 | 6.1 | 5 | 97.3 |
| PCPG | 179 | 148 | 84.6 | 20 | 11.4 | 6 | 3.4 | 4 | 97.8 |
| PRAD | 500 | 415 | 85.4 | 58 | 11.9 | 12 | 2.5 | 14 | 97.2 |
| READ | 172 | 82 | 92.1 | 6 | 6.7 | 1 | 1.1 | 83 | 51.7 |
| SARC | 261 | 228 | 90.5 | 18 | 7.1 | 6 | 2.4 | 9 | 96.6 |
| SKCM | 470 | 446 | 97.2 | 1 | 0.2 | 12 | 2.6 | 11 | 97.7 |
| STAD | 443 | 278 | 73.0 | 13 | 3.4 | 89 | 23.4 | 62 | 86.0 |
| TGCT | 263 | 119 | 92.2 | 6 | 4.7 | 4 | 3.1 | 134 | 49.0 |
| THCA | 507 | 334 | 80.7 | 27 | 6.5 | 52 | 12.6 | 93 | 81.7 |
| THYM | 124 | 103 | 84.4 | 6 | 4.9 | 13 | 10.7 | 2 | 98.4 |
| UCEC | 560 | 374 | 72.5 | 109 | 21.1 | 20 | 3.9 | 44 | 92.1 |
| UCS | 57 | 44 | 78.6 | 9 | 16.1 | 3 | 5.4 | 1 | 98.2 |
| UVM | 80 | 55 | 100.0 | 0 | 0.0 | 0 | 0.0 | 25 | 68.8 |

## Appendix E Zero-Score query Breakdown

Of the 440 benchmark queries, 17 received a score of zero. Their distribution by cause is given in Table VIII. No zero-score query produced a reversed or otherwise wrong analytical conclusion; the losses come from numerical divergence beyond tolerance, from reaching the turn limit, or from a formatting mismatch. The largest group is median survival, where the agent’s KM median (computed from raw survival times with censoring) diverges from the CDR’s tabulated median by more than the 5-month tolerance. The next group is filtered survival rates, where the set of patients with a given field in the raw files differs from the CDR patient set by more than the 5% tolerance. Three Tier 5 ranking queries reached the 12-turn limit before completing the full 33-type analysis, and one Tier 2 distribution query was scored zero due to a formatting mismatch between the reported distribution and the expected form.

**TABLE VIII.** Causes of the 17 zero-score queries.

| Cause | Count |
| --- | --- |
| Median survival divergence (KM vs CDR, >5 months) | 9 |
| Filtered survival coverage difference (>5%) | 4 |
| Tier 5 ranking reached 12-turn limit | 3 |
| Distribution formatting mismatch | 1 |
| **Total** | **17** |

## Appendix F

### Field Coverage Beyond the Curated Reference

Table IX lists the main clinical fields that the raw extraction recovers but the CDR does not contain, together with the number of patients for whom each field was extracted. These fields are the reason the extraction layer exists and are the basis for the paper’s claim that the extraction layer’s value lies in information the curated reference does not provide. For completeness we also note the four fields the CDR provides that the raw clinical files do not expose in the same curated form: cause of death, a curated clinical stage, and the PFI event and time.

**TABLE IX.** Clinical fields recovered by raw extraction that are absent from the curated CDR, with patient counts.

| Field | Patients |
| --- | --- |
| Drug treatments with dates and therapy type | 4,240 |
| Residual tumor status | 4,447 |
| Pathologic T stage | 7,888 |
| Pathologic N stage | 7,840 |
| Pathologic M stage | 6,968 |
| Lymph nodes positive count | 3,950 |
| Tobacco use history | 2,927 |
| Biomarker status (ER/PR/HER2; EGFR/KRAS) | cancer-specific |
| Biospecimen metadata (nuclei %, necrosis %) | cancer-specific |

## Appendix G Clinical Completeness Scoring Rules

The clinical completeness score is computed by pattern matching on the text of the agent’s response. Each component below is marked satisfied if any of its associated patterns is present. The score is the number of satisfied components divided by the number applicable to the query type, expressed as a percentage. Six components apply to every query; two additional components apply only to comparative queries. Table X lists each component and the textual cues used to detect it. The matching is deliberately simple and transparent so that scores are reproducible; it is not a substitute for expert human judgment of clinical quality, which is left to future validation.

**TABLE X.** Clinical completeness components and the textual cues used to detect each. components 7 and 8 are scored only for comparative queries.

| Component | Detected when the response contains |
| --- | --- |
| 1. Sample size reported | an explicit count such as “n = 412” or a phrase like “412 patients” |
| 2. Raw counts with rates | counts of alive and dead patients accompanying a rate, for example “230 alive, 180 dead” |
| 3. Proper survival methodology | survival-analysis language such as Kaplan-Meier, median survival, time-to-event, censoring, log-rank, or month-specific survival |
| 4. Data quality acknowledged | mention of missing data, coverage, incompleteness, small samples, or an explicit caution or caveat |
| 5. Contextualized statistics | a percentage paired with its underlying counts, or a count expressed as a fraction, for example “56% (230/410)” |
| 6. Source cited | reference to the data source (TCGA, CDR, raw extraction) |
| 7. Comparison direction (comparative only) | directional language such as higher, lower, greater, better, worse, older, or younger |
| 8. Statistical significance (comparative only) | mention of a p-value, a significance test (t-test, Mann-Whitney, chi-square, log-rank), or a stated significance level |

## Appendix H Numerical Accuracy Scoring Rules

The numerical accuracy scorer assigns each response a score in [0, 1] using rules specific to the query type. All rules operate by extracting numbers from the response text and comparing them against the CDR-derived ground truth within a tolerance. The rules are as follows.

### Single-patient lookups

Each requested field (age, gender, vital status) is checked independently, and the score is the fraction of fields reported correctly. Age must match exactly; gender and vital status are matched case-insensitively.

### Cohort and filtered survival rates

The scorer first tries to recover alive and dead counts from the response and, if their sum exceeds ten, computes the implied rate; a match within *±* 5 percentage points scores 1. Failing that, it searches for an explicitly stated survival percentage within *±* 5 points, then for any percentage within *±* 5 points, and finally for an event-count computation (patients minus events over patients). A 12-month KM value near the target receives partial credit of 0.5. Otherwise the score is 0.

### Cohort and filtered means (age)

Any number in the response within *±* 3 years of the ground-truth mean age scores 1; otherwise 0.

### Median survival

Any number within *±* 5 months of the ground-truth median scores 1; otherwise 0.

### Rankings

The score is 1 if the correct cancer-type code appears in the response, and 0 otherwise.

### Distributions

The score is the fraction of the three most frequent ground-truth categories that appear in the response.

### Comparative queries

These use weighted partial credit that sums to 1 when all elements are satisfied, as detailed in Table XI. Group identification contributes up to 0.3, each compared numerical value (survival rate within *±* 5 points, or mean age within *±* 3 years) contributes 0.15, a reported sample size per group contributes 0.05, and stating the correct direction of the comparison contributes 0.2. The final score is normalized by the total achievable weight for that query and capped at 1.

**TABLE XI.** Weights used by the comparative-query accuracy scorer. The score is the sum of achieved weights divided by the sum of applicable weights, capped at 1.

| Element | Weight |
| --- | --- |
| All groups named in the response | 0.30 |
| (partial: some but not all groups named) | 0.15 |
| Each compared value within tolerance (per value) | 0.15 |
| survival-rate tolerance | ±5 points |
| mean-age tolerance | ±3 years |
| Sample size reported (per group) | 0.05 |
| Correct direction of comparison stated (two-group) | 0.20 |

## Acknowledgment

During the preparation of this work the authors used an AI assistant [29] to improve the readability and language of portions of the Introduction, Discussion, and Conclusion. After using this tool, the authors reviewed and edited the content as needed and take full responsibility for the content of the publication. No external funding was received for this work. The authors declare no competing financial interests or personal relationships that could have appeared to influence the work reported in this paper.

## Data Availability

TCGA data are publicly available through the GDC Data Portal (https://portal.gdc.cancer.gov/). The CDR is available as supplemental material to Liu et al. [3]. The TCGA-Agent-Bench queries with their CDR-derived ground truth, the evaluation and scoring code, and the extraction pipeline are available from the corresponding author on reasonable request. The raw extracted patient records are derived from controlled and open TCGA sources through the GDC and are not redistributed directly.

---

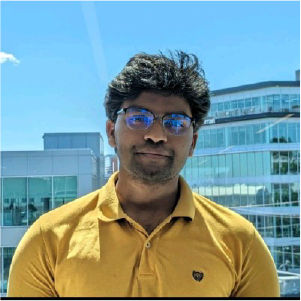

**RAJASHEKAR KORUTLA** is currently pursuing the Ph.D. degree in bioengineering with Northeastern University, Boston, MA, USA. His research focuses on building prediction models for healthcare outcomes.

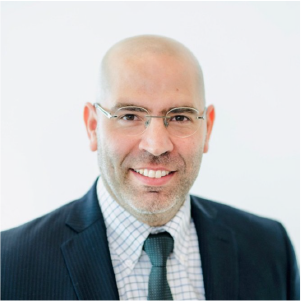

**SAEED AMAL** is currently an Assistant Research Professor with the Department of Bioengineering, Northeastern University, Boston, MA, USA. His research interests include artificial intelligence for healthcare, machine learning, natural language processing, and medical image processing.

## References

[1] J. N. Weinstein, E. A. Collisson, G. B. Mills, et al., “The Cancer Genome Atlas Pan-Cancer analysis project,” Nature Genet., vol. 45, pp. 1113–1120, 2013.

[2] K. Tomczak, P. Czerwinska, and M. Wiznerowicz, “The Cancer Genome Atlas (TCGA): An immeasurable source of knowledge,” Contemp. Oncol., vol. 19, no. 1A, pp. A68–A77, 2015.

[3] J. Liu, T. Lichtenberg, K. A. Hoadley, et al., “An integrated TCGA Pan-Cancer clinical data resource to drive high-quality survival outcome analytics,” Cell, vol. 173, no. 2, pp. 400–416, 2018.

[4] K. Singhal, S. Azizi, T. Tu, et al., “Large language models encode clinical knowledge,” Nature, vol. 620, pp. 172–180, 2023.

[5] L. Wang, C. Ma, X. Feng, et al., “A survey on large language model based autonomous agents,” Front. Comput. Sci., vol. 18, art. no. 186345, 2024.

[6] Z. Xi, W. Chen, X. Guo, et al., “The rise and potential of large language model based agents: A survey,” arXiv:2309.07864, 2023.

[7] S. Yao, J. Zhao, D. Yu, et al., “ReAct: Synergizing reasoning and acting in language models,” arXiv:2210.03629, 2023.

[8] Y. Zhu, P. Qiu, and Y. Ji, “TCGA-Assembler: Open-source software for retrieving and processing TCGA data,” Nature Methods, vol. 11, no. 6, pp. 599–600, 2014.

[9] A. Colaprico, T. C. Silva, C. Olsen, et al., “TCGAbiolinks: An R/Bioconductor package for integrative analysis of TCGA data,” Nucleic Acids Res., vol. 44, no. 8, art. no. e71, 2016.

[10] T. C. Silva, A. Colaprico, C. Olsen, et al., “TCGA Workflow: Analyze cancer genomics and epigenomics data using Bioconductor packages,” F1000Research, vol. 5, art. no. 1542, 2016.

[11] M. Cline, B. Craft, T. Swatloski, et al., “Exploring TCGA Pan-Cancer data at the UCSC Cancer Genomics Browser,” Sci. Rep., vol. 3, art. no. 2652, 2013.

[12] E. Cerami, J. Gao, U. Dogrusoz, et al., “The cBio Cancer Genomics Portal: An open platform for exploring multidimensional cancer genomics data,” Cancer Discovery, vol. 2, no. 5, pp. 401–404, 2012.

[13] J. Lee, W. Yoon, S. Kim, et al., “BioBERT: A pre-trained biomedical language representation model for biomedical text mining,” Bioinformatics, vol. 36, no. 4, pp. 1234–1240, 2020.

[14] Y. Gu, R. Tinn, H. Cheng, et al., “Domain-specific language model pretraining for biomedical natural language processing,” arXiv:2007.15779, 2021.

[15] S. M. Meystre, G. K. Savova, K. C. Kipper-Schuler, and J. F. Hurdle, “Extracting information from textual documents in the electronic health record: A review of recent research,” Yearbook Med. Informat., pp. 128–144, 2008.

[16] P. B. Jensen, L. J. Jensen, and S. Brunak, “Mining electronic health records: Towards better research applications and clinical care,” Nature Rev. Genet., vol. 13, no. 6, pp. 395–405, 2012.

[17] E. Alsentzer, J. Murphy, W. Boag, et al., “Publicly available clinical BERT embeddings,” in Proc. 2nd Clin. Natural Lang. Process. Workshop, 2019, pp. 72–78.

[18] J. Kefeli and N. Tatonetti, “Benchmark pathology report text corpus with cancer type classification,” medRxiv, 2023.

[19] J. Kefeli, J. Berkowitz, J. M. Acitores Cortina, et al., “Generalizable and automated classification of TNM stage from pathology reports with external validation,” Nature Commun., vol. 15, art. no. 8916, 2024.

[20] C. E. Jimenez, J. Yang, A. Wettig, et al., “SWE-bench: Can language models resolve real-world GitHub issues?” arXiv:2310.06770, 2024.

[21] C. Lu, C. Lu, R. T. Lange, et al., “The AI Scientist: Towards fully automated open-ended scientific discovery,” arXiv:2408.06292, 2024.

[22] J. Li, B. Hui, G. Qu, et al., “Can LLM already serve as a database interface? A big bench for large-scale database grounded text-to-SQLs,” arXiv:2305.03111, 2023.

[23] T. Schick, J. Dwivedi-Yu, R. Dessi, et al., “Toolformer: Language models can teach themselves to use tools,” in Adv. Neural Inf. Process. Syst., vol. 36, 2023.

[24] S. G. Patil, T. Zhang, X. Wang, and J. E. Gonzalez, “Gorilla: Large language model connected with massive APIs,” in Adv. Neural Inf. Process. Syst., vol. 37, 2024.

[25] Q. Jin, Y. Yang, Q. Chen, and Z. Lu, “GeneGPT: Augmenting large language models with domain tools for improved access to biomedical information,” Bioinformatics, vol. 40, no. 2, art. no. btae075, 2024.

[26] J. Linares-Blanco, A. Pazos, and C. Fernandez-Lozano, “Machine learning analysis of TCGA cancer data,” PeerJ Comput. Sci., vol. 7, art. no. e584, 2021.

[27] M. Mohammed, H. Mwambi, I. B. Mboya, et al., “A stacking ensemble deep learning approach to cancer type classification based on TCGA data,” Sci. Rep., vol. 11, art. no. 15626, 2021.

[28] A. Thennavan, F. Beca, Y. Xia, et al., “Molecular analysis of TCGA breast cancer histologic types,” Cell Genomics, vol. 1, no. 3, art. no. 100067, 2021.

[29] Anthropic, “The Claude API,” 2024. [Online]. Available: https://www.anthropic.com

